# Self-collected Saline Gargle Samples as an Alternative to Healthcare Worker Collected Nasopharyngeal Swabs for COVID-19 Diagnosis in Outpatients

**DOI:** 10.1101/2020.09.13.20188334

**Authors:** David M. Goldfarb, Peter Tilley, Ghada N. Al-Rawahi, Jocelyn A. Srigley, Geoffrey Ford, Heather Pedersen, Abhilasha Pabbi, Stephanie Hannam-Clark, Marthe Charles, Michelle Dittrick, Vijay J. Gadkar, Jeffrey M. Pernica, Linda M. N. Hoang

## Abstract

**Background:** We assessed the performance, stability, and user acceptability of swab-independent self-collected saliva and saline mouth rinse/gargle sample types for the molecular detection of SARS-CoV-2 in adults and school-aged children.

**Methods:** Outpatients who had recently been diagnosed with COVID-19 or were presenting with suspected COVID-19 were asked to have a nasopharyngeal swab collected and provide at least one self-collected sample type. A portion of participants were also asked about sample acceptability. Samples underwent molecular testing using multiple assays. Saline mouth rinse/gargle and saliva samples were tested daily at time zero, day one, and day 2 to assess nucleic acid stability at room temperature.

**Results:** 50 participants (aged 4 to 71 years) were included; of these, 40 had at least one positive sample and were included in the primary sample yield analysis. Saline mouth rinse/gargle samples had a sensitivity of 98% (39/40) while saliva samples had a sensitivity of 79% (26/33). Both saline mouth rinse/gargle and saliva samples showed stable viral RNA detection after 2 days of room temperature storage. Mouth rinse/gargle samples had the highest (mean 4.9) and HCW-collected NP swabs had the lowest acceptability scores (mean 3.1).

**Conclusion:** Saline mouth rinse/gargle samples demonstrated the highest combined user acceptability ratings and analytical performance when compared with saliva and HCW collected NP swabs. This sample type is a promising swab-independent option, particularly for outpatient self-collection in adults and school aged children.

## Background

Severe Acute Respiratory Syndrome Coronavirus 2 (SARS-CoV-2) has caused a global pandemic that has shown the potential to overwhelm the capacity of local and national health care systems (1). Rapid scale-up of molecular testing for SARS-CoV-2 has occurred on an unprecedented scale and has presented numerous challenges including deployment of assays with variable performance, lack of availability of high throughput testing platforms, and bottle-necks in supply chain procurement of pre-analytical materials and testing reagents (2). One of the key weaknesses in the diagnostic cycle has been the challenge of sample acquisition. While nasopharyngeal flocked swab (NPFS) collection has been the gold-standard for diagnosis of active SARS-CoV-2 infections using NAT methods (3), it is difficult to collect due to the associated discomfort, and is resource intensive requiring health care workers (HCW) for collection and procurement of these collection devices has proved to be a challenge in many jurisdictions. This quality initiative was carried out in order to meet anticipated increases in outpatient testing demands the expected challenges associated with global NPFS supply chain, the difficult nature of NPFS sample collection and complex handling of highly viscose saliva samples in the laboratory. We compared the analytical performance, sample stability and user acceptability of saline gargle and saliva samples against healthcare provider collected NPFS in out-patients with COVID-19 infections.

## Methods

Individuals from the Vancouver Greater Metropolitan Area of British Columbia were approached for participation in this quality-improvement initiative if they had SARS-CoV-2 detected in any clinical sample collected at an outpatient testing centre or if they were identified as a symptomatic household contact of a confirmed COVID-19 case. After August 28^th^, 2020 symptomatic children 4 to 12 years of age presenting to the BC Children’s and Women’s Hospital Campus COVID-19 Collection Centre were also asked to provide a saline mouth rinse/gargle sample in addition to an NPFS. Verbal consent was obtained from all participants and for those who provided both saliva and saline mouth rinse/gargle samples the order of sample collection was alternated sequentially (i.e. saliva first vs mouth rinse/gargle sample first). Demographic and clinical information was collected including age, whether they were a health care worker (HCW), if they had been previously admitted to hospital, the date of onset of their symptoms, and the date of initial molecular diagnosis of COVID-19 (if applicable). Sample collection occurred either at the participant’s residence or at the BC Children’s and Women’s Hospital Campus COVID-19 Collection Centre, and instruction sheets (**Supplement 1**) with supporting verbal instructions were provided. Participants were asked to not eat, drink, smoke, brush their teeth or chew gum 1 hour prior to collection.

### Self-collection procedures

Nasopharyngeal flocked swabs (flexible flocked swabs with 3 mL universal viral transport system media, Beckon Dickinson, Sparks, MD, USA) were collected via the left naris unless preference was stated for the right naris. NPFS were inserted the distance from the naris to the external ear canal and then rotated 5 times and left in place for 5 – 10 seconds prior to being removed as per Center for Disease Control instructions for collection (4). Mouth rinse/gargle specimens were self-collected by instructing users to open 5 ml vials of sterile 0.9% saline (Addipak®, Teleflex Medical, Research Triangle Park, NC, USA) and squeeze the contents into their or their child’s open mouth. They were then asked to swish the contents for 5 seconds followed by tilting their heads back and gargling for 5 seconds. This swish/gargle cycle was repeated 2 more times and then the saline was expelled into a wide mouthed sterile empty polypropylene container (Leakbuster™ 90 ml container, Starplex Scientific, Etobicoke, Ontario, Canada). For saliva sample collection, participants were asked to pool and spit saliva repeatedly into a wide mouthed sterile empty polypropylene container (Leakbuster™ 90 ml container, Starplex Scientific, Etobicoke, Ontario, Canada) until at least 5 – 10 ml were collected if possible. Samples were immediately brought to the laboratory and were processed within 12 hours of collection. All samples were vortexed for at least 10 seconds prior to aliquoting for testing.

### Laboratory testing

All samples were extracted with the QiaSymphony automated extractor using the DSP virus/pathogen minikit (Qiagen, Germantown, MD, USA) and subsequently tested with a laboratory developed test (LDT) within 24 hours of collection on the Applied Biosystems 7500 Fast Real-Time PCR System (Life Technologies, Carlsbad, CA). Mouth rinse/gargle and saliva samples were also stored in the laboratory at room temperature (21°C) and extracted and tested again on day 1 and day 2 after collection. The previously validated LDT is a triplex reverse transcription polymerase chain reaction (RT-PCR) assay that targets both the pan-sarbecovirus E gene as well as the RNA-dependent RNA polymerase (RdRP) gene and also targets the human RNase P gene as a sample positive control (5). A portion of the NPFS and mouth rinse gargle specimens were also tested using the Cepheid Xpert Xpress SARS-CoV-2 assay on the GeneXpert system (Cepheid, Sunnyvale, CA) as per manufacturer instructions.

### User acceptability of sampling

Participants who had all three samples collected were asked about the acceptability of each self-collected sample type they had obtained as well as about the HCW-collected NPFS using a 5 point Likert scale with 1 being “least acceptable” and 5 being “most acceptable”.

This project was reviewed by the BC Children’s and Women’s Research Ethics Board and deemed a quality improvement/quality assurance (QI/QA) activity based on completion of the Provincial Health Services Authority Project Sorting Tool.

### Statistical analysis

For the purpose of the analysis of sensitivity for each sample type, individuals were considered a positive case if they had at least one sample test positive for both targets (E gene and RdRP gene) on the LDT. Samples were considered positive with a threshold cycle (Ct) value of less than 40 on the LDT and as reported by the Xpert assay (presumptive positive results were considered as positive). Using this reference standard for a SARS-CoV-2 positive participant the sensitivity results of mouth rinse/gargle sampling was compared to NPFS and saliva sampling (LDT assay) using the test for comparing two independent proportions (STATA command prtesti); the McNemar exact test was also used to determine the comparability of mouth rinse/gargle sample testing as compared to NPFS or saliva sample testing. The Ct values obtained with baseline testing using all sample types were compared using repeated-measures ANOVA using Box’s conservative correction factor, with *post hoc* significant pairwise differences determined using Tukey’s HSD; this methodology was also used to compare acceptability ratings of all sample types. For assessment of stability of SARS-CoV-2 RNA between gargle samples and saliva samples (transport media free methods), individuals were only included if they had samples tested at each of those three time points (time zero, day 1, and day 2). Statistical significance was set as p<0.05. All testing was done using STATA v16.1 (College Station, TX).

## Results

A total of 50 participants (34 known positives and 16 household contacts/symptomatic children) provided validation samples between May 8^th^ and September 11^th^, 2020. Of these, 40 participants were found to be positive according to the reference definition at the time of study sample collection. There were 28 (56%) participants that were female, with a median age of 25.1 years (25-75 percentile 13.6-35.9 years) and 10 were < 18 years of age. Data on symptoms was available for 40 participants; of these, 17 (42%) had fever and 25 (62%) had cough. Among participants found to be positive, the median time from infection confirmation to re-testing was 3 days (25-75 percentile 2-5 days) and the median time from symptoms to re-testing was 7 days (25-75 percentile 4-9 days).

The results of testing for each of the sample types for all positive participants are shown in **Table 1**. Mouth rinse/gargle samples were significantly more likely to be positive than saliva samples (difference of 18.7%, 95%CI 3.9-33.5%, p=0.01). When matched samples were compared, mouth rinse/gargle sample testing results differed statistically from saliva samples (26 positive by both, 6 rinse/gargle positive but saliva negative, and 1 negative by both, McNemar p=0.03) but mouth rinse/gargle sample and saliva testing results were statistically similar to HCW-collected NPFS testing results. Positivity proportions for NPFS, mouth rinse/gargle and saliva sample types based on time from COVID-19 diagnosis are shown in **Figure 1**.

**Table 1.** Performance of sample types in 40 COVID-19 positive participants.

| Study ID | NP Swab (LDT) | NP Swab (Gx) | Gargle (LDT) | Gargle (Gx) | Saliva (LDT ) |
| --- | --- | --- | --- | --- | --- |
| 1 | P | P | P | P | P |
| 2 | P | P | P | P | N |
| 3 | N | PP | P | NT | N |
| 4 | P | P | P | PP | N |
| 5 | P | P | P | P | P |
| 6 | NT | NT | P | P | P |
| 7 | P | P | P | P | P |
| 8 | P | P | P | P | P |
| 9 | P | P | P | P | P |
| 10 | P | P | P | P | N |
| 11 | P | P | P | P | P |
| 12 | P | P | P | P | P |
| 13 | P | P | P | P | P |
| 14 | P | P | P | P | P |
| 15 | P | P | P | P | P |
| 16 | P | P | P | P | P |
| 17 | P | P | P | P | P |
| 18 | P | P | P | P | P |
| 19 | P | P | P | P | P |
| 20 | NT | NT | P | P | P |
| 21 | NT | NT | P | P | P |
| 22 | P | P | P | P | P |
| 23 | P | P | P | P | P |
| 24 | P | P | P | P | N |
| 25 | P | P | P | P | P |
| 26 | P | P | P | P | P |
| 27 | P | P | P | P | P |
| 28 | N | N | P | P | P |
| 29 | P | P | P | P | P |
| 30 | P | P | P | N | P |
| 31 | P | P | N | P | N |
| 32 | P | P | P | P | N |
| 33 | P | P | P | P | P |
| 34 | P | NT | P | P | NT |
| 35 | P | NT | P | P | NT |
| 36 | P | NT | P | P | NT |
| 37 | P | NT | P | P | NT |
| 38 | P | NT | P | P | NT |
| 39 | P | NT | P | P | NT |
| 40 | P | NT | P | P | NT |
| N=40 | 35/37 | 29/30 | 39/40 | 38/39 | 26/33 |
| Sensitivity | NA | NA | 97.5% | 97.4% | 78.8% |
| 95% CI | NA | NA | 86.8, 99.9 | 86.5, 99.9 | 61.0, 91.0 |

NP = nasopharyngeal, LDT = laboratory developed test, Gx = genexpert assay, NA = not applicable, P = positive test, N = negative test, PP = presumptive positive, NT = not tested, CI = confidence interval

**Figure 1.**
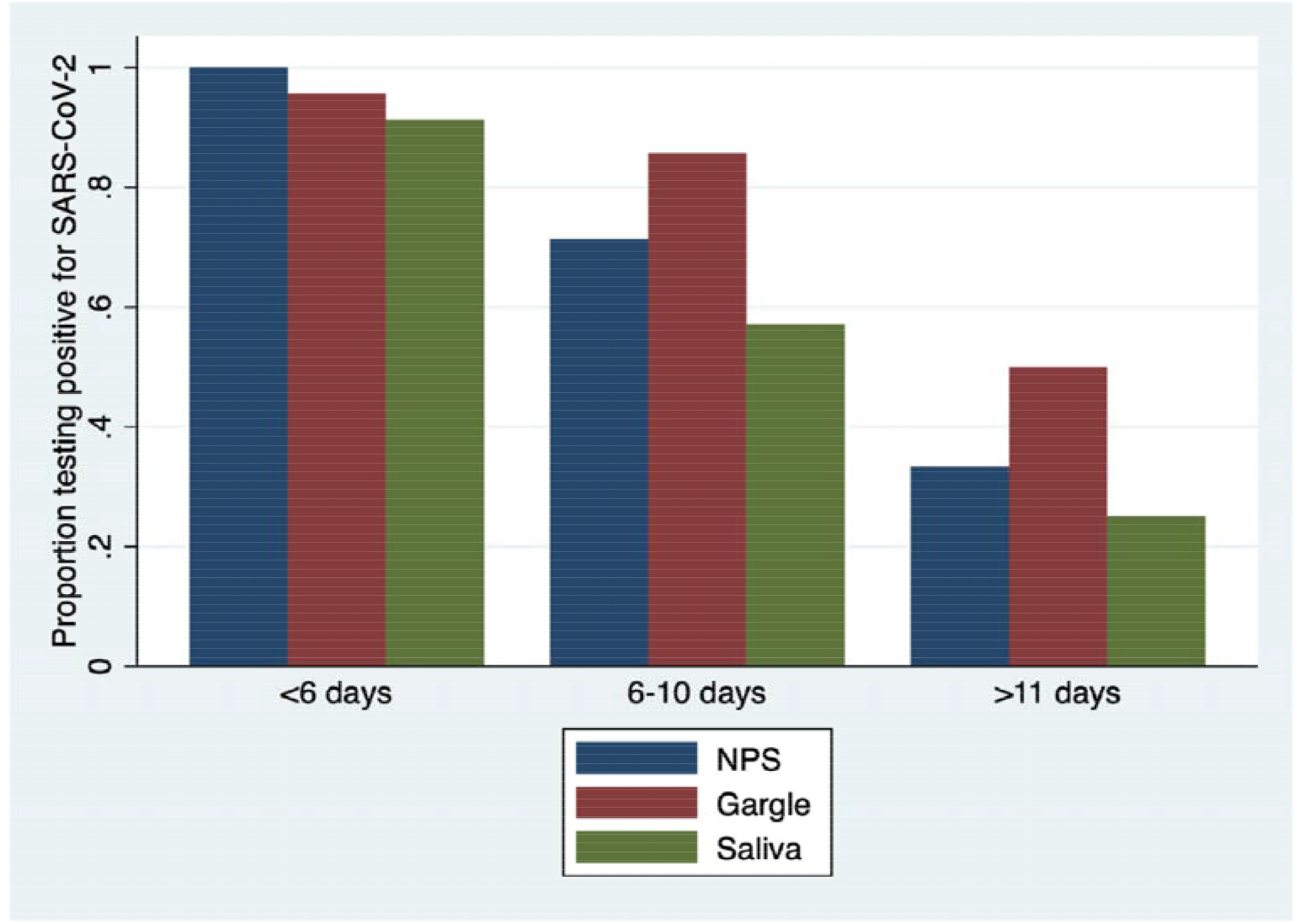
Proportion of samples testing positive based on time from COVID-19 diagnosis.

Participant rated acceptability performance of sample types is shown in **Table 2**. There were clear differences between the acceptability of the sample types; the mouth rinse/gargle sample had the highest mean acceptability (4.95) and was significantly more acceptable than HCW-collected NPFS (mean acceptability 3.17) or saliva sampling (mean acceptability 4.44).

**Table 2.** Acceptability of sample types as rated on a 5 point Likert scale (1= lowest acceptability and 5 = highest acceptability)

| Sample | Mean acceptability | Acceptability difference |  | 95%CI of difference | Tukey p of difference |
| --- | --- | --- | --- | --- | --- |
| Mouth Rinse/Gargle | 4.95 | vs: saliva NPFS | 0.50<br>1.78 | 0.0074 to 0.99<br>1.28 to 2.27 | 0.046<br><0.001 |
| Saliva | 4.44 | vs: NPFS | 1.28 | 0.78 to 1.77 | <0.001 |
| NPFS | 3.17 |  |  |  |  |

NPFS = nasopharyngeal flocculated swab, CI = confidence interval

Mean Ct values at baseline did not differ significantly between sample types; these data are shown in **Figure 2**. Mean Ct data at day zero, day 1, and day 2 for mouth rinse/gargle and saliva samples are shown in **Figure 3**. There was no significant difference in baseline Ct values across sample types, and for saliva and mouth rinse/gargle samples there was no significant loss of RNA recovery over all time points. Also amongst positive participants with mouth rinse/gargle (n = 30) and saliva (n = 28) samples that were tested on all three days there were a similar number of negative results (for both targets) on each day (saliva day 0=6, day 1 = 4 and day 2 = 3; mouth rinse/gargle day 0 = 1, day 1 = 5, and day 2 = 2).

**Figure 2:**
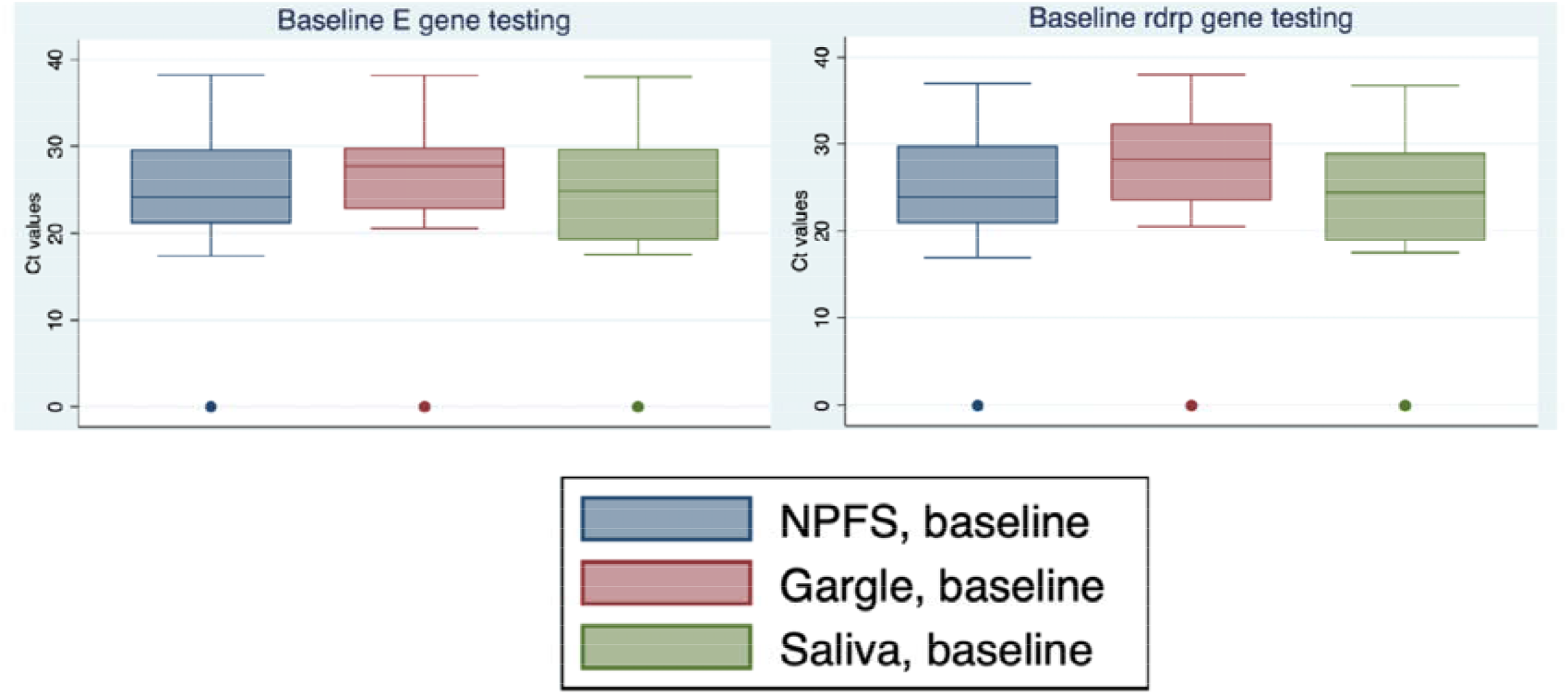
Mean threshold cycle (Ct) values across sample types at baseline.

**Figure 3.**
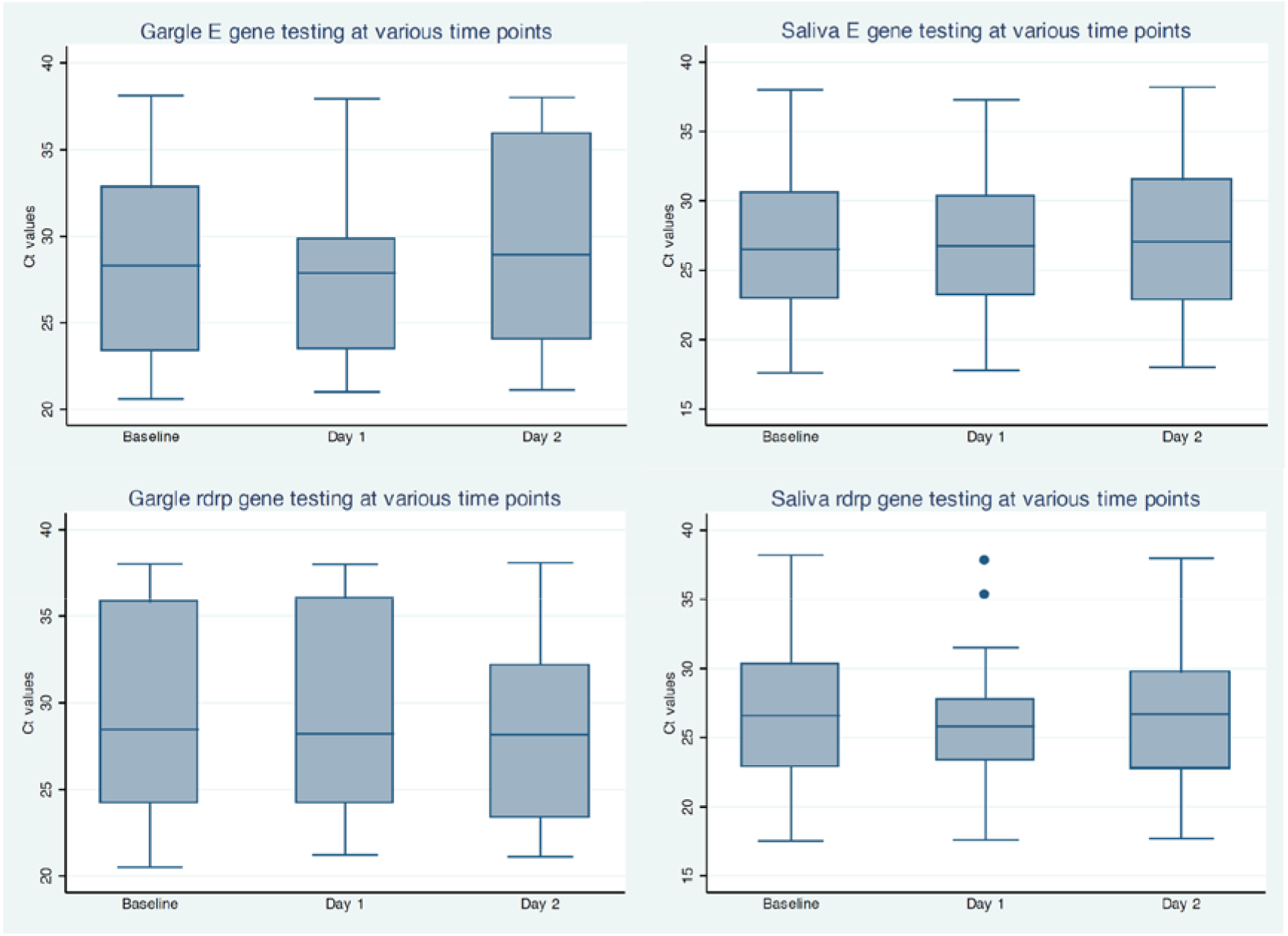
Mean threshold cycle (Ct) values at baseline, Day 1, and Day 2 for gargle and saliva samples with positive results.

## Discussion

Laboratory diagnostics are critical for any attempt to contain the COVID-19 pandemic. Our study demonstrates that self-collected mouth rinse/gargle samples are non-inferior to HCW-collected NPFS for the detection of SARS-CoV-2 and, at the same time, are significantly more acceptable to patients. Mouth rinse/gargle specimens, in particular, demonstrated the highest sensitivity and were preferred by those undergoing sampling. These data should be considered by all those planning laboratory testing strategies and algorithms; self-collected mouth rinse/gargle samples may obviate the need for the deployment of significant numbers of HCWs trained in sample collection and the consumption of large amounts of personal protective equipment. It should be emphasized that testing volumes will very likely be even higher in late 2020 as many children and adolescents return to school and respiratory symptoms become more common with the increasing prevalence of many other respiratory viruses.

Saline mouth rinse/gargle samples (also known as mouth throat washes) have previously been evaluated for influenza and other respiratory virus detection (6, 7), showing promising performance when compared to throat swab and other sample types. Interestingly, although there have been numerous evaluations of multiple self-collected sample types including mid-turbinate swabs, throat swabs, and saliva samples, there have been very few reports to date describing mouth rinse/gargle samples for COVID-19 diagnosis (8, 9). In one study in Germany (10), 5 individuals were evaluated who had both saline gargle samples and throat swab samples and all 5 gargle samples were positive whereas only 4 had a positive throat swab sample. The technique we used for acquisition of saline mouth rinse/gargle samples (three cycles for mouth rinse followed by gargle) may allow for improved recovery of viral RNA given that it would effectively sample the entire oropharynx. Collecting mouth rinse/gargle samples could also potentially result in a significant amount of savings due to the lack of need for personal protective equipment or trained HCWs for sample acquisition. Its utility might be even higher in low and middle income country settings or remote regions where access to testing clinics would be another barrier to sample acquisition. The stability of RNA recovery in this sample type was preserved for at least two days at room temperature, making later drop-off delivery of samples a feasible option.

In our study, saliva samples were both significantly less sensitive and less acceptable than mouth rinse/gargle samples. Several recent reports have found favourable detection rates for saliva samples, however most of these have been in inpatient populations (11) where viral load levels tend to be higher and/or utilized additional manual saliva processing steps (12) that are less amenable to high throughput processing. In our evaluation processing of saliva samples was intermittently observed to be impeded by variable sample viscosity; additional manual processing of two saliva samples was required due to excessive amounts of mucus present. For these reasons, amongst the swab- and transport media free options, the mouth rinse/gargle samples appeared more attractive.

Our study design had several strengths. In addition to having a number of participants across the pediatric and adult age ranges, we also enrolled solely individuals presenting with outpatient illness, where the largest burden of testing is performed. This is particularly the case for testing surges that are expected with return to school for children in the fall. Our evaluation also includes the assessment of performance across multiple extraction and PCR platforms, including a Health Canada and FDA authorized commercial assay. A main weakness of the study is that a number of the patients enrolled did not have samples collected until > 5 days into their illness and therefore we had a number of samples with lower amounts of viral RNA detected. Also as mentioned, the population sampled only included relatively well outpatients and it is unclear if similar performance would be found in those with more severe inpatient disease.

Given the very high user acceptability rating, lack of need for swabs and/or transport media, and excellent diagnostic yield, saline mouth rinse/gargle samples appear to be a preferred sample type for testing of outpatients with suspected COVID-19.

## Data Availability

The authors confirm that the data supporting the findings of this study are available within the article [and/or] its supplementary materials.

## Acknowledgements

We would like to thank the participants who agreed to contribute to this validation project and also the nursing staff at the BC Children’s and Women’s Campus COVID-19 Collection Centre and the molecular microbiology technologists at the BC Children’s Hospital and the BC Centre for Disease Control for their contribution to this work.

